# Population-Scale Liver Volume Nomograms: Data-Driven Insights from High-Throughput Imaging and Clinical Data Integration

**DOI:** 10.1101/2025.07.17.25331751

**Authors:** Yuqi Wang, Marilyn Yamamoto, Mark Martin, Jacob A. Macdonald, Diana Kadi, Kyle J. Lafata, Mustafa R. Bashir

## Abstract

**Background:** Liver volume is a potentially useful biomarker for clinical and research applications. However, comprehensive population-scale understanding of the association between liver volume and disease states is lacking.

**Purpose:** To identify key factors influencing liver volume in a large and diverse cohort using a data-driven method.

**Materials and Methods:** This retrospective study included 78,983 patients who underwent abdominal CT and MR imaging within a single health system between 2014 and August 2024. Images were processed using an automated pipeline that employed a nnUNet model for liver segmentation and a ResNet-based quality control module. Clinical data, including demographic, physiological, diagnosis codes, and lab results within three months of imaging were integrated. Feature selection utilized LASSO regression, and predictive models were validated across a reserved test set and a temporal validation cohort.

**Results:** Eight variables associated with liver volume were identified: body surface area (BSA), weight, age, liver steatosis, race (Black or African American), smoking status, nicotine dependence, cigarettes (uncomplicated), and fever (unspecified). Nomograms derived from these predictors achieved root mean squared errors (RMSE) of 374 mL to 413 mL (Z-score 0.70–0.78) and correlation coefficients (R^2^) of 0.41–0.44 across datasets. Adjusted models incorporating diagnostic codes and lab results consistently outperformed demographic-only models.

**Conclusion:** Our findings determined the association between patient characteristics and liver volume, and establish nomograms that may be applicable across populations. The identification of these factors advances our understanding of the drivers of liver volume.

**Summary:** Our study leverages high-throughput analysis of imaging and clinical data to identify key predictors of liver volume, providing robust nomograms for accurate prediction and improving understanding of liver volumetry.

**Key Results:**

1. From analysis of 145,165 abdominal scans, we identified eight key predictors of liver volume, with BSA (668 mL), liver steatosis (+268 mL), and unspecified fever (+150 mL) showing the strongest associations.
2. A model incorporating these factors had moderate performance for predicting liver volume (R2=0.41-0.44, RMSE=374.07-405.7) in test sets.
3. A simplified model based on five “easily observable” factors had similar performance for predicting liver volume (R^2^, RMSE) in the same test sets.

## INTRODUCTION

Liver volume serves as a valuable biomarker providing insights in clinical decision-making. Among its established applications are treatment planning for liver malignancies [1, 2], prognostic evaluations for hepatocellular carcinoma, assessing future liver remnant in liver resections [3, 4], and guiding living donor liver transplantation. Liver volume is also vital in prognostic evaluations for conditions such as acute liver failure and Fontan-associated liver diseases, as well as in managing Gaucher’s disease [5, 6], and other related conditions. Emerging applications include its use in monitoring nonalcoholic steatohepatitis (NASH)/metabolic associated steatohepatitis (MASH) and treatment follow-up [7-9].

Despite its significance, liver volumetry has primarily been confined to specific, narrow contexts, leaving its broader potential to uncover population-wide patterns and associations largely unexplored. This limited scope is partly due to the labor-intensive nature of traditional manual measurements, which has made large-scale analyses impractical [10-12]. Though some researchers like Small et al. [13] have attempted to expand the application of liver volumetry, comprehensive studies examining its relationships with diverse health and disease states across large, heterogeneous populations are still lacking.

Recent advances in deep learning-based automated segmentation algorithms, combined with CT and MR imaging, have made large-scale liver volume measurements feasible [14-16]. While several studies have leveraged these technologies, they have focused on specific clinical scenarios. Kim et al. [17] established reference intervals for liver volume using a cohort of 3,461 healthy adult liver donors and 158 viral hepatitis patients. Perez et al. [18] analyzed 3,065 patients to develop a weight-based threshold for hepatomegaly, while Choi et al. [19] investigated the relationship between hepatic steatosis and liver volume in 1,038 healthy adult liver donors. However, these studies have not yet fully explored the potential broader relationships between liver volume and various clinical factors across the general patient population. Current understanding of what influences liver volume is still mainly limited to basic metrics, such as body weight, and a few specific disease conditions.

The purpose of our study was to assess liver volumes across a large, diverse patient population. Our hypothesis was that there would be previously unidentified associations between liver volume and clinical and demographic factors.

## MATERIALS AND METHODS

### Study Patients

This study was approved by the *[BLINDED FOR REVIEW]* Institutional Review Board (IRB) for retrospective data analysis. As shown in Fig.1(a), the initial cohort comprised all patients undergoing abdominal axial CT and MR scans conducted in our health system between 2014 and August 2024. The cohort was split into three subsets: a development set using data from 2014 to 2021, a reserved test set with data from 2022 and allowing for patient overlap with the development set, and a temporal validation set with new patient data between 2023 and Aug 2024 and no patient overlap with the other two sets.

**Figure 1:**
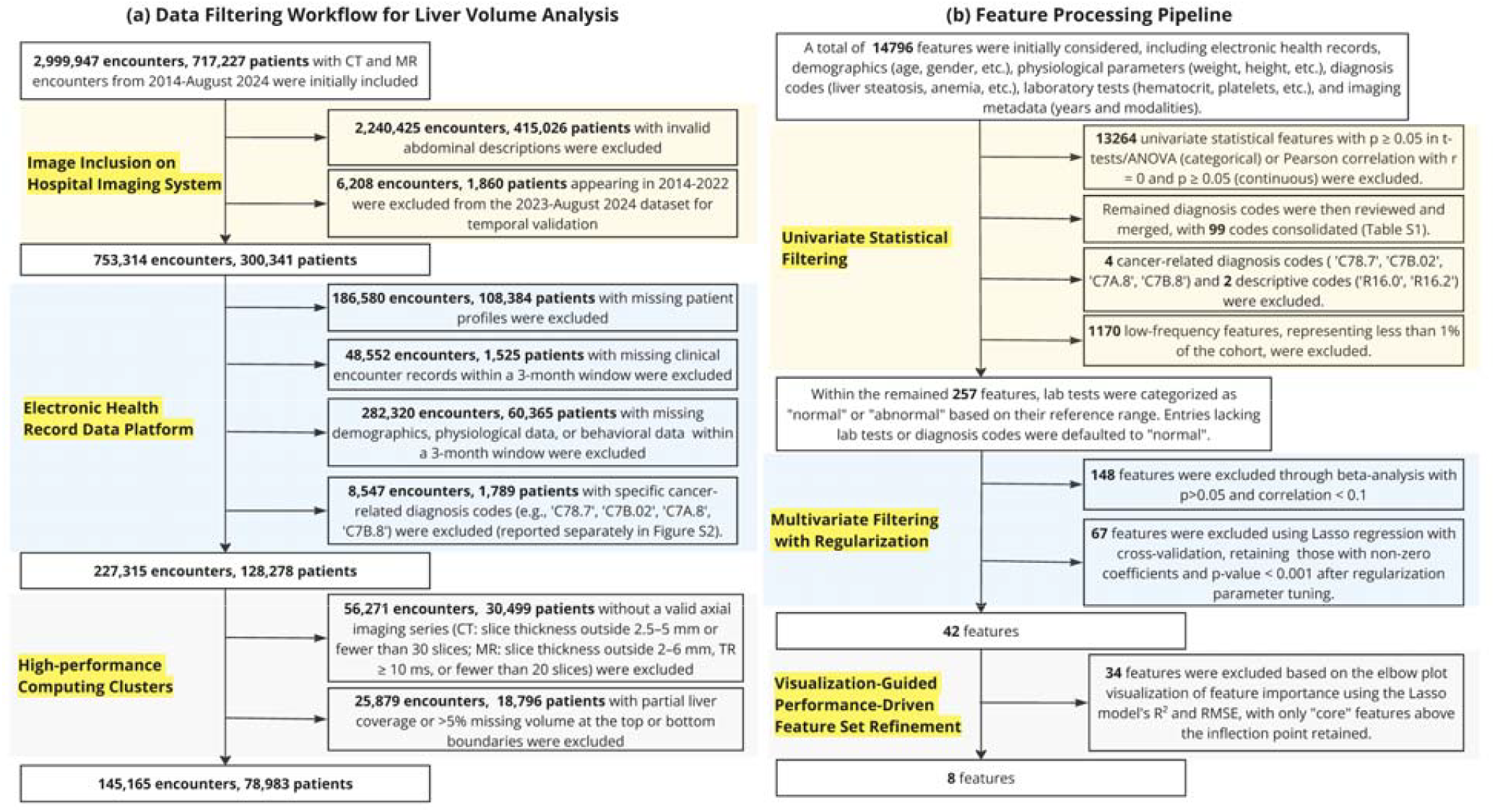
Data Filtering and Feature Selection Workflows. **(a)** Data filtering steps from 2,996,947 encounters and 717,227 patients initially to 145,165 encounters and 78,983 patients with inclusion/exclusion criteria. **(b)** Feature selection pipeline reducing 14,796 initial features to 8 final predictors through univariate statistical filtering, multivariate filtering with regularization, and visualization-guided refinement.

### Cohort Selection

From the initial cohort, encounters with invalid abdominal study descriptions were excluded. Further filtering on our electronic health record (EHR) platform excluded encounters missing patient profiles, clinical records, demographics, physiological data, or behavioral data within a three-month window. Encounters with specific cancer-related diagnosis codes (i.e., ‘C78.7’, ‘C7B.02’, ‘C7A.8’, ‘C7B.8’) were excluded (Fig. S2).

On our high-performance computing clusters, the dataset was further refined based on imaging features, in MRI particularly to identify fat-suppressed T1-weighted images. Encounters without a valid axial imaging series (CT slice thickness outside 2.5-5mm or <30 slices; MR slice thickness outside 2-6mm, TR > 10 ms, or <20 slices) were excluded. When multiple valid series were available per patient, MR series were prioritized based on keywords (“portal”, “pre”, “upper”, and “dixon w”); while for CT the first valid series was selected. If no valid series was identified based on these rules, the examination was discarded. Imaging equipment was from two vendors (GE Healthcare, Waukeshaw, WI, USA; Siemens Healthineers, Forscheim/Erlangen, Germany) available in our institution’s clinical image archive.

### Automated High-throughput Liver Volume Collection

Liver volumes were obtained using a high-throughput pipeline (Fig. S1). After identifying a single series for analysis, liver segmentation was performed using a previously reported nnUNet model [20, 21] using Nvidia RTX A6000 GPUs. A ResNet-based edge-quality control module validated segmentation outputs, excluding cases where portions of the liver were excluded at the edges of the imaging volume (Appendix S1). Liver volumes were then calculated using the stack of cylindroids method [22]. The pipeline was developed in Python 3.7.10, using libraries including PyTorch 1.13.1+cu117 and Pandas 1.3.0, as well as shell scripts for workflow integration.

### EHR Data Collection

Non-imaging data included demographics, physiological parameters, diagnosis codes, and laboratory results within three months of imaging. Demographic data included age, gender, ethnicity, race (first race recorded for multi-racial patients), and smoking status (self-reported). Physiological parameters included height, weight, and body surface area (BSA) calculated using the Mosteller equation [23].

Diagnosis codes, recorded in ICD-9 and ICD-10 formats, were standardized by converting ICD-9 codes to ICD-10. To enhance data interpretability, clinically similar ICD-10 codes were merged based on their likely relevance to liver volume and reviewed by a physician (*[initials blinded for review]*; Table S1). We focused on associations that may not be immediately clinically evident based on patient history, and so excluded patients with specific cancer-related diagnosis codes (e.g., ‘C78.7’, ‘C7B.02’, ‘C7A.8’, ‘C7B.8’), reported in Appendix Fig. S2. This was based on the assumption that the overall clinical outcomes in this patient group are driven by their malignancies rather than systemic factors affecting liver volume, as well as anecdotal observations that these patients can develop very enlarged livers due to hepatic metastatic disease. Descriptive diagnosis codes, including R16.0 (Hepatomegaly, not elsewhere classified) and R16.2 (Hepatomegaly with splenomegaly, not elsewhere classified), were also excluded as they were not expected to provide information about specific disease states and conceptually overlap with the quantitative volume measurements.

Laboratory test results were classified as “normal” or “abnormal” based on their respective laboratory-specific reference ranges. Following the principle of Documentation by Exception, where only abnormal findings are typically documented, absent tests or diagnosis codes within the three-month window were interpreted as no clinical suspicion for abnormality [24, 25]. This approach minimized data exclusion and allowed for a broader analysis rather than being restricted to a limited subset of complete records.

### Data-driven Feature Selection

Feature selection and model development were conducted exclusively on the development set to avoid data leakage and ensure unbiased performance evaluation. The initial feature set comprised EHR, demographics, physiological parameters, laboratory tests, and imaging metadata (years and modalities) to account for potential temporal trends and modality-specific variations in measurements.

As shown in Fig. 1(b), we employed a multi-stage feature refinement approach. First, univariate statistical analysis was used to remove variables failing to meet significance thresholds (p ≥ 0.05 in t-tests/ANOVA for categorical variables; or Pearson correlation with r = 0 and p ≥ 0.05 for continuous variables). The remaining diagnosis codes were then reviewed and semantically similar codes consolidated (Table S1), and low-frequency features (<1% of cohort) were removed for model stability.

Then, interdependencies between variables were considered and multivariate analysis with regularization applied. Beta analysis [26] was used to quantify both the magnitude and direction of feature associations with liver volume. To further select a robust feature set, we then employed Lasso regression [27] with cross-validation, applying L1 regularization to shrink less important feature coefficients to zero, effectively performing feature selection while preventing overfitting. For both beta analysis and Lasso regression, we retained only features with non-zero coefficients and p-value < 0.001 after tuning the regularization parameter.

Finally, we implemented a visualization-guided performance-driven approach to identify the most important variables for balance between model complexity and predictive power. An elbow plot illustrated feature importance via Lasso-derived R^2^ and root mean squared error (RMSE), with “core” features retained for the nomogram.

### Data Analysis

The nomogram was derived using core features from the development set. Variables were categorized as control variables (easily obtainable information such as patient age and gender) and non-control variables (less obvious values such as diagnosis codes and laboratory values). A baseline predictive model was developed using linear regression with weighted mean squared error (MSE) loss using control variables. Residual adjustments were then modeled incrementally using non-control variables, keeping control variable coefficients fixed. Model performance was tested on both the reserved test and the temporal validation sets. To enhance interpretability and facilitate standardized comparisons across different scales, we implemented parallel analyses using z-scores. All data analysis was performed in Python 3.7.10 using libraries including Pandas 1.3.0, Scikit-learn 1.0.2, and Scipy 1.7.0.

## RESULTS

### Dataset Characteristics

After applying the inclusion/exclusion criteria illustrated in Fig. 1(a), the complete study dataset consisted of 78,983 unique patients and 145,165 total examinations (126,079 CT and 19,805 MR), with 54,656 unique patients and 98,271 examinations allocated to the development set, 26,455 unique patients and 38,149 examinations to the reserved test set, and 6,019 unique patients and 8,745 examinations to the temporal validation set. Mean age was 58 ± 17 years with 52.4% (76,039/145,165) females. Demographic and biophysical parameters are summarized in Table 1. The measured range of liver volumes was 1696.0 ± 532.46 mL. A histogram summary of liver volumes, as well as comparison between individuals without any positive diagnosis codes or abnormal lab results and the general population are shown in Fig. 2.

**Table 1:**
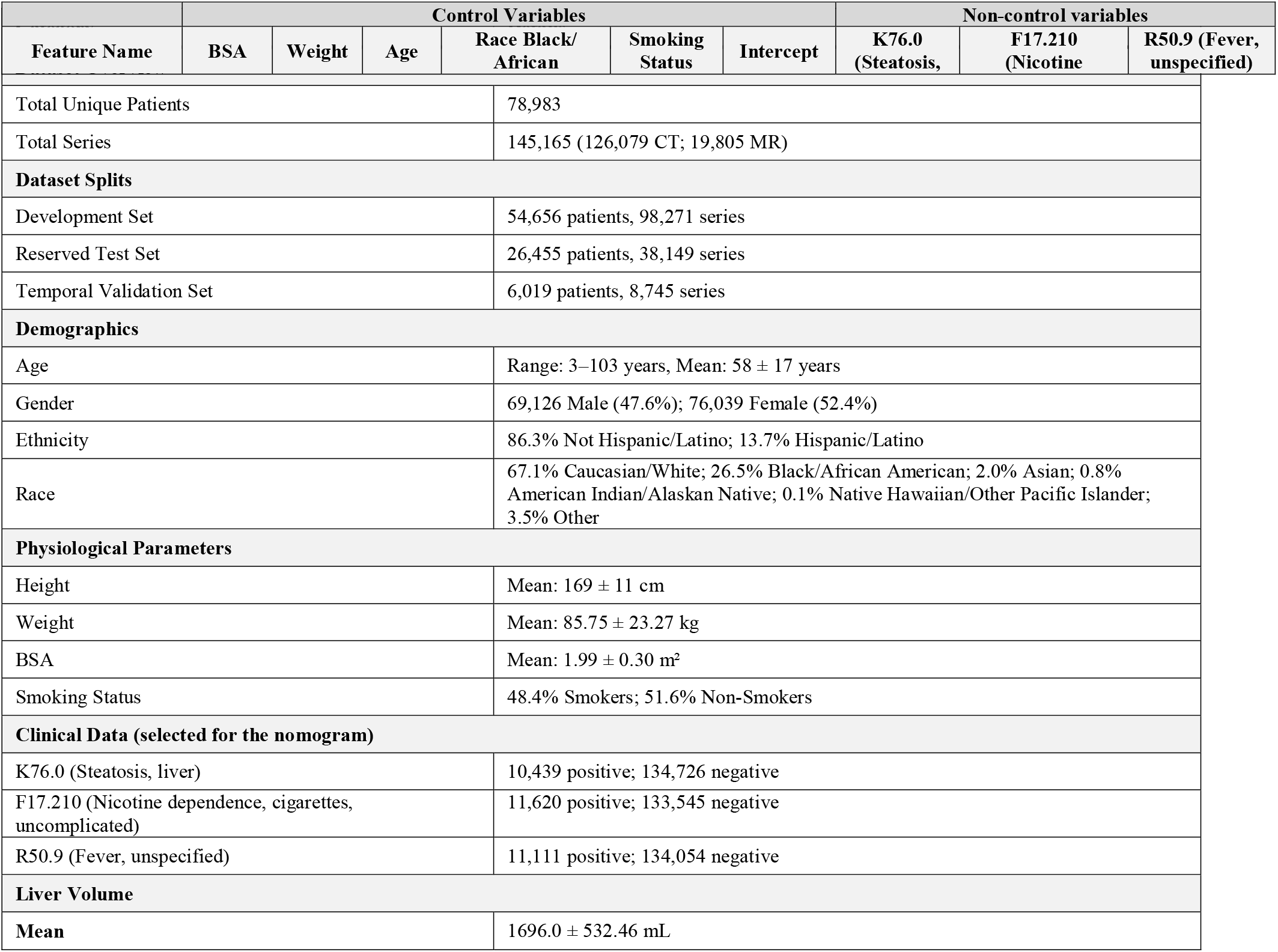
Dataset Characteristics.

**Figure 2:**
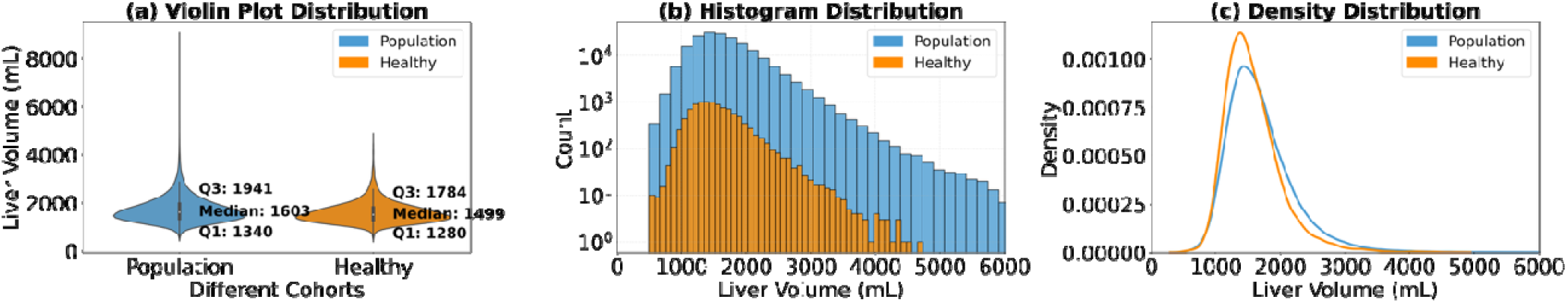
Comparison of distribution statistics across two cohorts: Population, and Healthy. **(a)** Violin plots showing the distribution of values with medians and interquartile ranges for each group. **(b)** Log-scale histograms illustrating the frequency distribution of values across the groups. **(c)** Kernel densit estimation (KDE) plots highlighting the probability density for each group.

### Feature Selection

As shown in Fig. 1(b), after initially considering a total of 14,796 features, the univariate and multivariate analyses identified 42 potentially relevant features. After ranking feature importance using elbow point analysis (Fig. 3), a final set of eight “core” features was selected, including BSA, weight, age, liver steatosis, race (Black or African American), smoking status, nicotine dependence, cigarettes (uncomplicated) and fever (unspecified) (Fig. 3). BSA, weight, race, and smoking status were designated as control variables. The eight core features were then used for subsequent nomogram development and evaluation.

**Figure 3:**
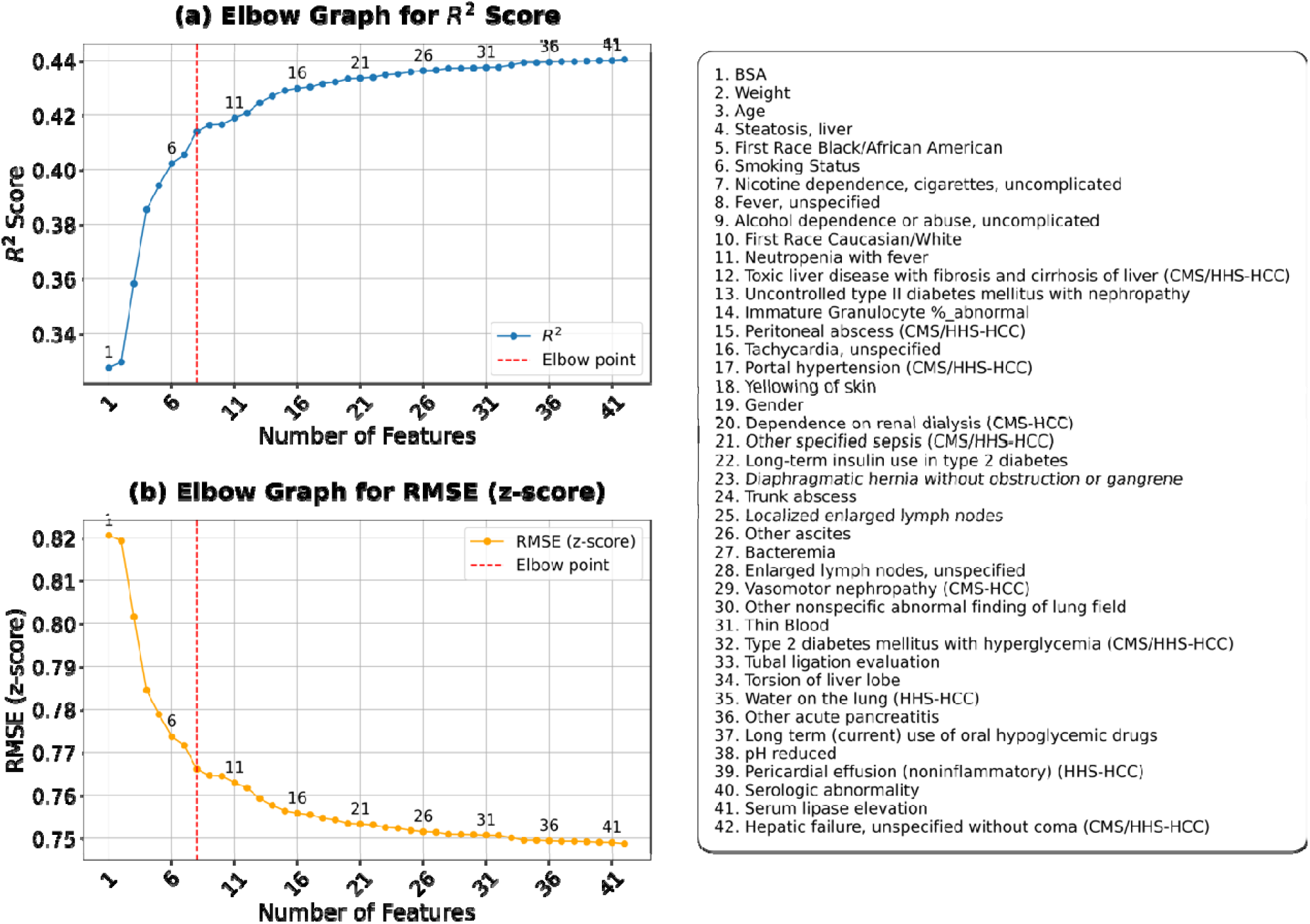
Feature selection analysis using R^2^ and root mean squared error (RMSE) metrics to identify the optimal number of features for the model. **(a)** and **(b)** plot R^2^ values and RMSE (z-score) values against the number of features, with the elbow point (feature #8) marked, emphasizing the optimal feature count to minimize error.

### Nomogram and Performance

The derived nomogram (Table 2) demonstrated the relative contribution of each feature to liver volume prediction, and Table 3 shows a comparison between a baseline model using only control variables to an adjusted model incorporating all eight core variables. Performance metrics (Table 3) indicated a modest improvement from the baseline to adjusted model. While the absolute gains were small (a 3% increase in R^2^ and a 2–3% reduction in RMSE), the consistency of improvement across all datasets suggested that the non-control variables add meaningful predictive value to the nomogram.

**Table 2:**
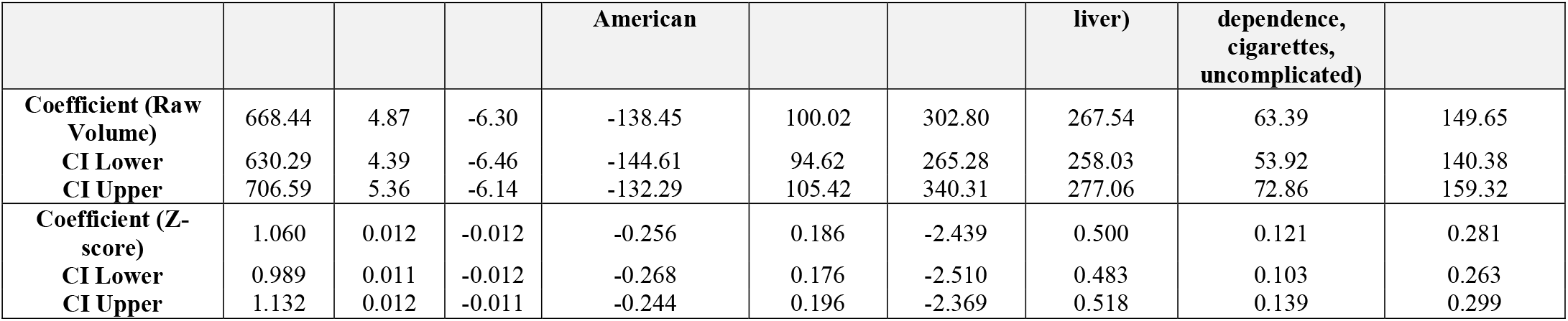
Derived Nomograms.

**Table 3:** Statistical Nomogram Performance.

|  | Baseline Model |  |  | Adjust Model |  |  |
| --- | --- | --- | --- | --- | --- | --- |
|  | R<sup>2</sup> | RMSE | RMSE (Z-score) | R<sup>2</sup> | RMSE | RMSE (Z-score) |
| Developing Set | 0.38 | 423.49 | 0.80 | 0.41 | 413.4 | 0.78 |
| Reserved Test Set | 0.38 | 415.2 | 0.78 | 0.41 | 405.7 | 0.76 |
| Temporal Validation Set | 0.41 | 381.44 | 0.72 | 0.44 | 374.07 | 0.70 |

To understand the effects of individual features, Figs. 4 and 5 illustrate their impact on liver volume. Regression plots of continuous variables (Fig. 4) highlighted BSA as the strongest correlate (coefficient: 668.44, 95% CI: 630.29–706.59). Both BSA and weight showed a positive correlation with liver volume, while age exhibited a weak negative correlation. Violin plots (Fig. 5) illustrate the impact of categorical variables, with liver steatosis showing the largest effect. Fever, smoking status, and nicotine dependence were associated with slightly increased liver volumes, while race was associated with a slight decrease.

**Figure 4:**
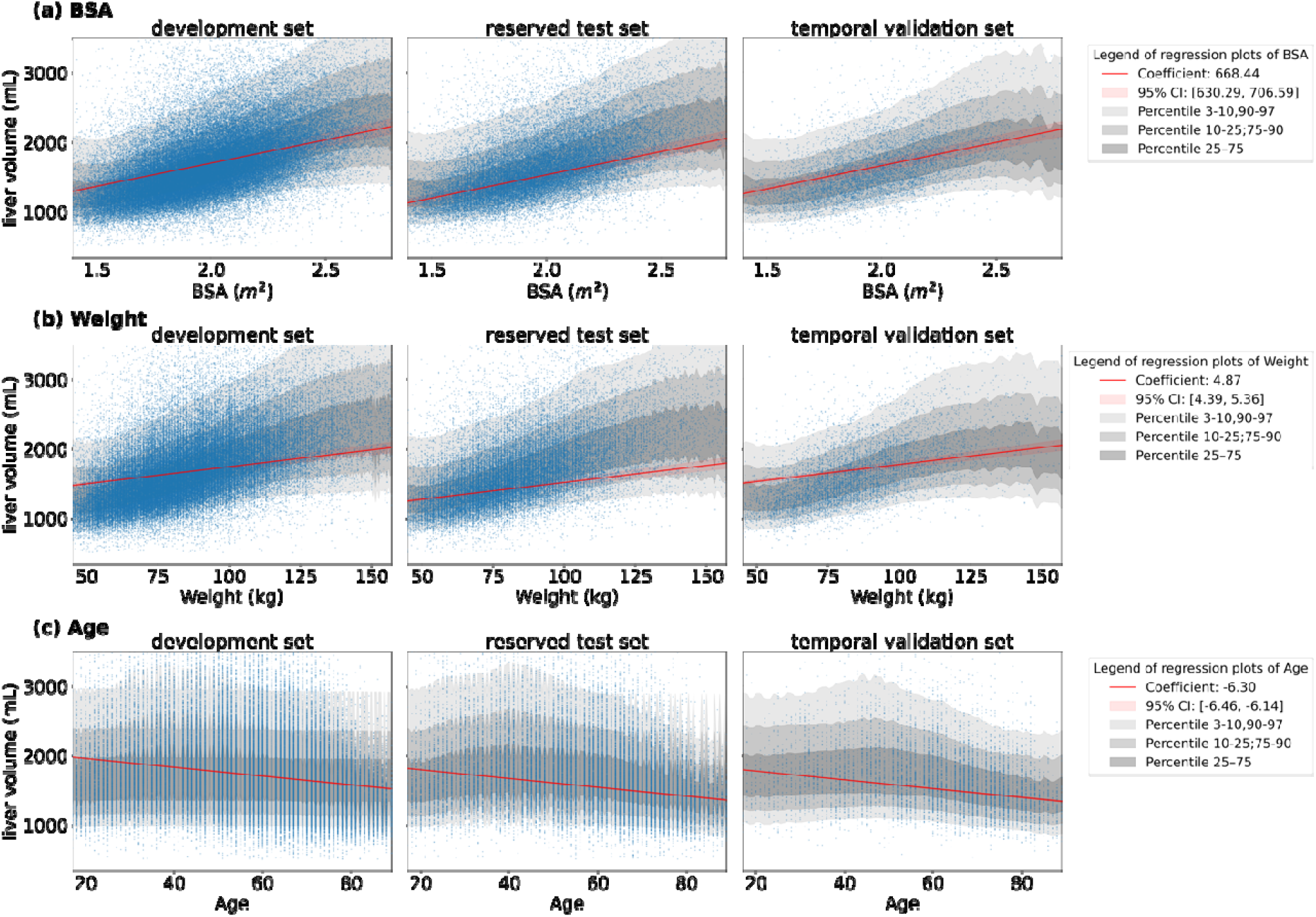
Regression analysis plots of key continuous features with coefficient confidence intervals and percentiles. **(a) (b)** and **(c)** are regression plots showing the correlation between liver volume and body surface area (BSA), weight, and age, respectively. From left to right columns, they display the regression on the development set, reserved test set, and temporal validation set respectively. The red line represents the regression line, with pink shaded regions depicting coefficient confidence intervals (95%). Percentile bands (3-10, 25-75, 10-90, and 90-97) are shaded in different gray scales to highlight data distribution.

**Figure 5:**
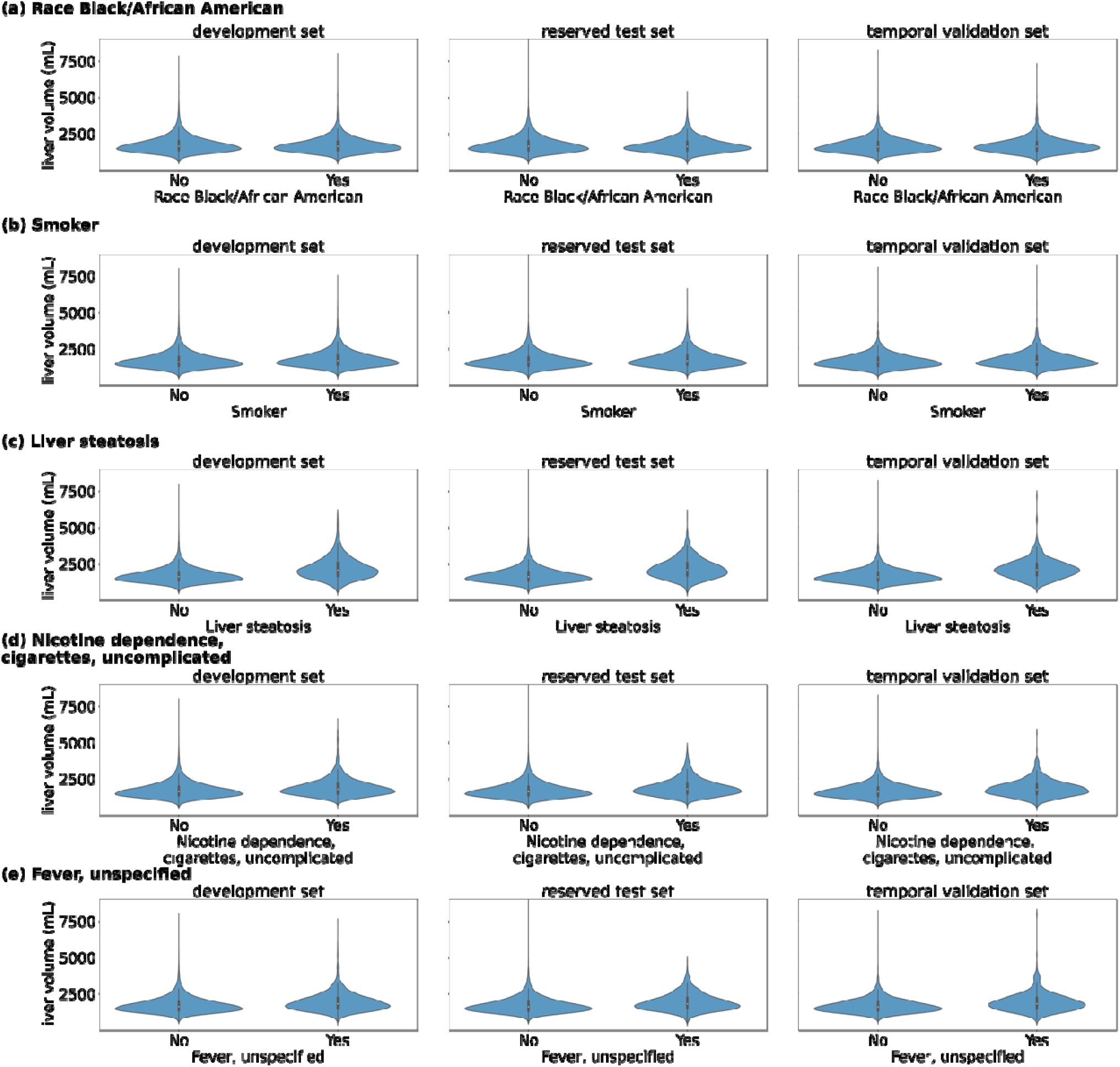
Violin plots illustrating the distribution of key categorical variables for positive and negative classes across the three datasets. Panels **(a), (b), (c), (d)**, and **(e)** represent the race, smoking status, liver steatosis, nicotine dependence, and fever, respectively. From left to right, the plots display the distributions for the development set, reserved test set, and temporal validation set. The violin shapes represent the kernel density estimation of the data distribution, with the white dot marking the median, and the black bar indicating the interquartile range.

Finally, to illustrate the progressive changes in model fit as variables were added, Fig. 6 presents density plots. In Fig. 6, panels (a), (b), and (c) illustrate the alignment between predicted and true liver volumes as continuous variables are added. Panel (d) demonstrated the closest alignment between predicted and true liver volumes across all datasets with all variables. The inclusion of categorical variables slightly widened the distribution, improving the consideration of larger liver volumes while having a limited impact on the overall cohort due to the relatively small proportion of positive cases as shown in Table 1.

**Figure 6:**
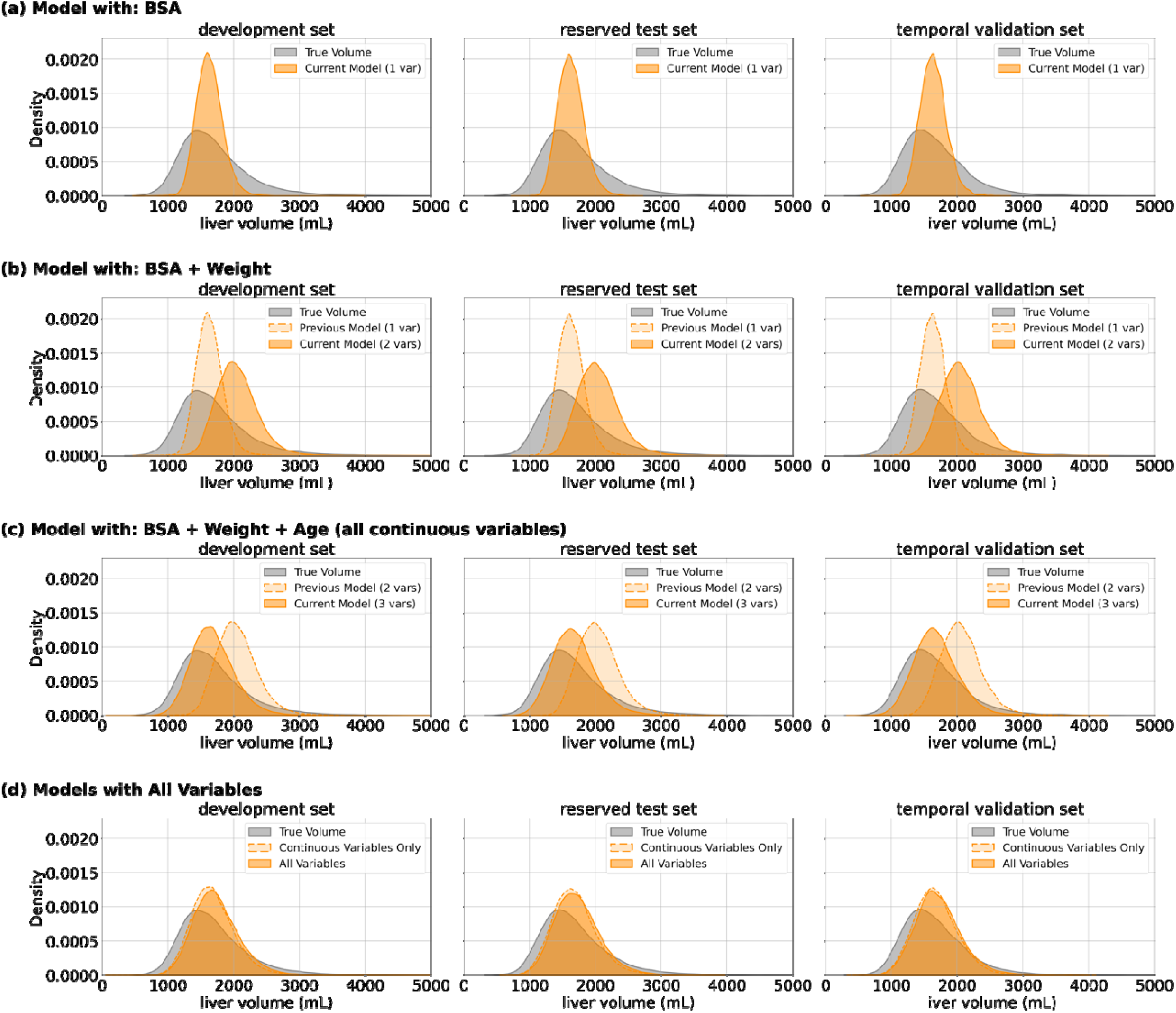
Density plots showing the progressive effect of adding different features on liver volume distribution across datasets. Distribution of true liver volumes (gray) versus predicted values (yellow) for (a) BSA-only model, (b) BSA+Weight model, (c) BSA+Weight+Age model, and (d) models with all variables (including all categorical features), demonstrating progressive improvement in alignment between predicted and actual distributions. Each panel contains three plots representing the development set, reserved test set, and future validation set from left to right. For clearer comparison, the model from the previous panel is also included in the current panel, shown in dashed line.

## DISCUSSION

Our study presents a data-driven approach to understanding factors influencing liver volume and demonstrates the relationship between clinical data and liver volume. By employing a high-throughput automated pipeline and leveraging both imaging and clinical data, we identified eight key factors associated with liver volume. Statistical nomograms incorporating these predictors achieved consistent performance across development (R^2^ = 0.41, RMSE = 413.4 mL), reserved testing (R^2^ = 0.41, RMSE = 405.7 mL), and temporal validation sets (R^2^ = 0.44, RMSE = 374.07 mL).

The strong correlations observed between liver volume and physiological parameters such as BSA, weight, and age are consistent with prior studies on liver volume determinants. These variables likely reflect both the metabolic demands and anthropometric variations that influence organ size. Clinical conditions showed substantial effects on liver volume, with liver steatosis associated with the largest increase (268 mL), followed by unspecified fever (150 mL) and uncomplicated nicotine dependence, cigarettes (63 mL). Demographic factors also demonstrated significant associations, with race Black/African American associated with a decrease of 138 mL and positive smoking status corresponding to an increase of 100 mL in liver volume. It should be noted that recent publications have suggested that race may in fact be a proxy for social determinants of health, rather than reflecting an association with the patient’s genetic origins [28-30]. Notably, despite including laboratory tests and imaging metadata (modality and year) in our initial feature set, our data-driven selection process did not identify these as significant correlates for liver volume. Nicotine dependence, identified through diagnostic codes, indicates a more severe and chronic exposure than self-reported smoking status, which could contribute to liver volume changes through long-term metabolic and inflammatory effects [31]. Unspecified fever, often described clinically as fever of unknown origin (FUO), has over 200 potential malignant, infectious, and inflammatory causes, with up to 51% of cases remaining undiagnosed despite extensive workup.

Non-clue-based CT imaging and other tests are frequently ordered, with a subset of such cases present in our dataset. FUO remains a challenging diagnosis in immunocompetent adults, and its association with increased liver volume may reflect underlying systemic effects warranting further investigation [32]. The incorporation of these clinical factors improved model performance only slightly.

This study introduces several advancements in liver volume analysis. First, we developed a high-throughput pipeline for automated liver segmentation and validation using deep learning models, enabling large-scale volumetric analysis across one of the most diverse cohorts reported to date. Importantly, we included an automated quality control module to remove imaging examinations in which the liver was only partially included, which are common in clinical practice and could artifactually influence the observed distribution of liver volumes. In contrast to previous studies constrained by smaller datasets (n=1038-3461 [17][18] [19]) our larger dataset (78,983 unique patients and 145,165 total imaging time points) enabled rigorous validation on both a reserved test set and a temporal validation cohort, ensuring broad generalizability across temporal and population variations. Second, while Kim et al. [17] focused on healthy donors and viral hepatitis patients, and Perez et al. [18] examined only healthy individuals undergoing colorectal screening or renal donor evaluation, our study encompasses a broader patient population with diverse, abnormal clinical conditions. This allows for more generalizable insights into liver volume variations across different health states. In addition, integrating clinical features such as diagnosis codes, and laboratory results and data-driven feature selection allowed us to identify key correlates of liver volume and develop nomograms for predicting liver volumes, enhancing both accuracy and clinical applicability. This approach provides insights into previously unexplained variations in liver volume, which were largely based on weight-focused studies by Perez et al. [18]or limited demographic variables (age, sex, height, and weight) with inconsistent association with liver volume (p-values ranging from <.001 to .73 across different percentiles) as observed by Kim et al [17]. In contrast, our model identified and validated a more comprehensive set of variables with robust association with liver volume. Furthermore, while Choi et al. [19] investigated the relationship between steatosis and liver volume in a healthy donor population, our study validates and extends these findings across a broader patient population, providing more generalizable insights into how steatosis affects liver volume in various clinical contexts.

Our work has limitations. First, each step of the analysis has vulnerabilities to error. For instance, the liver segmentation and quality control models are well validated but not perfectly accurate. Additionally, to enhance the dataset, we combined laboratory results and diagnosis codes recorded within three months of the imaging with the corresponding liver volumes. Values obtained further in time from the date of imaging examination may less accurately reflect the patient’s current state at the time of imaging. Also, although assuming normal values for missing laboratory and diagnostic data is a statistically accepted approach, it could introduce additional noise since the true state of those values was not known.

Additionally, the temporal association between diagnosis codes and imaging examinations is not well captured in our EHR data. Second, our dataset primarily consists of patients undergoing abdominal CT/MR imaging for clinical indications, which may not represent the general population not undergoing imaging. Additionally, while our models incorporate demographic and physiological features, other potentially relevant factors such as dietary habits, physical activity levels, etc. were not readily available.

To address these limitations, future studies should aim to further assess our findings in broader, more diverse populations, including healthy individuals without clinically-required abdominal CT/MR imaging, and others. Prospective studies integrating longitudinal imaging and clinical data could also provide deeper insights into dynamic changes in liver volume over time and their clinical implications.

Incorporating more diverse data types into predictive models may offer a more comprehensive understanding of liver volumetry. Moreover, while the current work relies on a linear nomogram for prediction, future efforts could investigate deep generative modeling approaches—such as conditional variational autoencoders—to capture complex, nonlinear relationships and enable personalized liver volume estimation with individualized reference intervals.

In conclusion, our study identified eight key factors associated with liver volume: BSA, weight, age, liver steatosis, race, smoking status, nicotine dependence, and fever. The implementation of a high-throughput automated pipeline enabled large-scale analysis across 14,165 encounters and 78,983 patients, revealing associations that were previously undetectable in smaller studies. These findings confirm our hypothesis that there are previously unidentified associations between liver volume and various clinical and demographic factors beyond the basic metrics previously established.

## Data Availability

Data generated or analyzed during the study are available upon reasonable request to the authors.

## Abbreviations

BSA: body surface area,
CI: confidence intervals,
MSE: mean squared error,
RMSE: root mean squared error,
EHR: electronic health record

