## Appendix for "Population-Scale Liver Volume Nomograms: Data-Driven Insights from High-Throughput Imaging and Clinical Data Integration"

Table S1: Diagnosis Codes Transformation

| Normalized Code | Diagnosis Code | Diagnosis Name | Diagnosis Code Type |
|---|---|---|---|
| A04.7 | A04.72 | Enterocolitis due to Clostridium difficile, not specified as recurrent | ICD-10-CM |
| | A04.7 | Enterocolitis due to Clostridium difficile | ICD-10-CM |
| A41.9 | 995.91 | Tracheostomy, sepsis | ICD-9-CM |
| | A41.89 | Other specified sepsis (CMS/HHS-HCC) | ICD-10-CM |
| | A41.9 | Sepsis, unspecified organism (CMS/HHS-HCC) | ICD-10-CM |
| B37.89 | B37.0 | Candidal stomatitis | ICD-10-CM |
| | B37.89 | Other sites of candidiasis | ICD-10-CM |
| | B37.9 | Candidiasis, unspecified | ICD-10-CM |
| B95.61 | B95.61 | Methicillin susceptible Staphylococcus aureus infection as the cause of diseases classified elsewhere | ICD-10-CM |
| | B95.62 | Methicillin resistant Staphylococcus aureus infection as the cause of diseases classified elsewhere | ICD-10-CM |
| C16.9 | C16.9 | Malignant neoplasm of stomach, unspecified (CMS/HHS-HCC) | ICD-10-CM |
| | 151.9 | Stomach cancers | ICD-9-CM |
| C34.10 | C34.12 | Malignant neoplasm of upper lobe, left bronchus or lung (CMS/HHS-HCC) | ICD-10-CM |
| | C34.11 | Malignant neoplasm of upper lobe, right bronchus or lung (CMS/HHS-HCC) | ICD-10-CM |
| | 162.3 | Syndromes, Pancoast's | ICD-9-CM |
| C34.90 | C34.90 | Malignant neoplasm of unspecified part of unspecified bronchus or lung (CMS/HHS-HCC) | ICD-10-CM |
| | 162.9 | Undifferentiated carcinoma of lung (CMS/HHS-HCC) | ICD-9-CM |
| | C34.91 | Malignant neoplasm of unspecified part of right bronchus or lung (CMS/HHS-HCC) | ICD-10-CM |
| | C34.92 | Malignant neoplasm of unspecified part of left bronchus or lung (CMS/HHS-HCC) | ICD-10-CM |
| C50.4 | C50.912 | Malignant neoplasm of unspecified site of left female breast (CMS/HHS-HCC) | ICD-10-CM |
| | 174.4 | Malignant neoplasm of upper-outer quadrant of female breast (CMS/HHS-HCC) | ICD-9-CM |
| | 174.8 | Malignant neoplasm of other specified sites of female breast (CMS/HHS-HCC) | ICD-9-CM |
| | C50.811 | Malignant neoplasm of overlapping sites of right female breast (CMS/HHS-HCC) | ICD-10-CM |
| | C50.911 | Malignant neoplasm of unspecified site of right female breast (CMS/HHS-HCC) | ICD-10-CM |
| | 174.9 | Squamous cell carcinoma, breast | ICD-9-CM |
| | C50.412 | Malignant neoplasm of upper-outer quadrant of left female breast (CMS/HHS-HCC) | ICD-10-CM |
| | C50.919 | Malignant neoplasm of unspecified site of unspecified female breast (CMS/HHS-HCC) | ICD-10-CM |
| | C50.812 | Malignant neoplasm of overlapping sites of left female breast (CMS/HHS-HCC) | ICD-10-CM |
| C61 | C61 | Malignant neoplasm of prostate (CMS/HHS-HCC) | ICD-10-CM |
| | 185 | Prostrate cancers | ICD-9-CM |
| C79.31 | C79.31 | Secondary malignant neoplasm of brain (CMS/HHS-HCC) | ICD-10-CM |
| | 198.3 | Secondary squamous cell carcinoma of brain (CMS/HHS-HCC) | ICD-9-CM |
| D50.0 | D53.9 | Nutritional anemia, unspecified | ICD-10-CM |
| | D50.0 | Iron deficiency anemia secondary to blood loss (chronic) | ICD-10-CM |
| | D50.9 | Iron deficiency anemia, unspecified | ICD-10-CM |
| | 285.29 | Iron reutilization anemia | ICD-9-CM |
| | 280.9 | Sideropenic anemia with reticuloendothelial siderosis | ICD-9-CM |
| D61.810 | 284.19 | Other pancytopenia (CMS/HHS-HCC) | ICD-9-CM |
| | D61.810 | Antineoplastic chemotherapy induced pancytopenia (CMS-HCC) | ICD-10-CM |
| | D61.818 | Other pancytopenia (CMS/HHS-HCC) | ICD-10-CM |
| D63.8 | D63.1 | Anemia in chronic kidney disease | ICD-10-CM |
| | D63.8 | Anemia in other chronic diseases classified elsewhere | ICD-10-CM |
| D62 | D62 | Acute posthemorrhagic anemia | ICD-10-CM |
| | 285.1 | Postoperative anemia due to acute blood loss | ICD-9-CM |
| D64.9 | D64.9 | Anemia, unspecified | ICD-10-CM |
| | D64.89 | Other specified anemias | ICD-10-CM |
| D68.4 | D68.4 | Acquired coagulation factor deficiency (CMS/HHS-HCC) | ICD-10-CM |
| | 286.9 | Warfarin-induced coagulopathy (CMS/HHS-HCC) | ICD-9-CM |
| D69.6 | 287.5 | Transient thrombocytopenia (CMS-HCC) | ICD-9-CM |
| | D69.6 | Thrombocytopenia, unspecified (CMS-HCC) | ICD-10-CM |
| | D69.59 | Other secondary thrombocytopenia | ICD-10-CM |
| D70.9 | D70.9 | Neutropenia, unspecified (CMS-HCC) | ICD-10-CM |
| | 780.61 | Neutropenia with fever | ICD-9-CM |
| D84.9 | D84.9 | Immunodeficiency, unspecified (CMS/HHS-HCC) | ICD-10-CM |
| | 279.9 | Unspecified disorder of immune mechanism (CMS/HHS-HCC) | ICD-9-CM |
| | D89.9 | Disorder involving the immune mechanism, unspecified (CMS/HHS-HCC) | ICD-10-CM |
| E11.42 | 357.2 | Visceral diabetic neuropathy | ICD-9-CM |
| | E11.42 | Type 2 diabetes mellitus with diabetic polyneuropathy (CMS/HHS-HCC) | ICD-10-CM |
| E11.65 | E11.65 | Type 2 diabetes mellitus with hyperglycemia (CMS/HHS-HCC) | ICD-10-CM |
| | E11.649 | Type 2 diabetes mellitus with hypoglycemia without coma (CMS/HHS-HCC) | ICD-10-CM |
| E11.8 | E11.8 | Type 2 diabetes mellitus with unspecified complications (CMS/HHS-HCC) | ICD-10-CM |
| | 250.02 | Uncontrolled type II diabetes mellitus with nephropathy | ICD-9-CM |

| | | | |
|---|---|---|---|
| E66.01 | E66.9 | Obesity, unspecified | ICD-10-CM |
| | 278.01 | Severely overweight | ICD-9-CM |
| | E66.01 | Morbid (severe) obesity due to excess calories (CMS/HHS-HCC) | ICD-10-CM |
| E86.1 | E86.1 | Hypovolemia | ICD-10-CM |
| | E86.9 | Volume depletion, unspecified | ICD-10-CM |
| E87.2 | 276.2 | pH reduced | ICD-9-CM |
| | E87.2 | Acidosis | ICD-10-CM |
| | E87.20 | Acidosis, unspecified | ICD-10-CM |
| E87.70 | 276.69 | Other fluid overload | ICD-9-CM |
| | E87.70 | Fluid overload, unspecified | ICD-10-CM |
| | E87.79 | Other fluid overload | ICD-10-CM |
| E87.5 | E87.5 | Hyperkalemia | ICD-10-CM |
| | E87.6 | Hypokalemia | ICD-10-CM |
| | 276.7 | Tacrolimus-induced hyperkalemia | ICD-9-CM |
| E87.0 | E87.0 | Hyperosmolality and hypernatremia | ICD-10-CM |
| | E87.1 | Hypo-osmolality and hyponatremia | ICD-10-CM |
| | 276.1 | Sodium, decreased level | ICD-9-CM |
| E83.51 | E83.51 | Hypocalcemia | ICD-10-CM |
| | E83.52 | Hypercalcemia | ICD-10-CM |
| F10.20 | F10.20 | Alcohol dependence, uncomplicated (CMS/HHS-HCC) | ICD-10-CM |
| | F10.10 | Alcohol abuse, uncomplicated | ICD-10-CM |
| F11.90 | F11.20 | Opioid dependence, uncomplicated (CMS/HHS-HCC) | ICD-10-CM |
| | F11.90 | Opioid use, unspecified, uncomplicated | ICD-10-CM |
| F32.8 | 311 | Unipolar Depressions | ICD-9-CM |
| | F43.21 | Adjustment disorder with depressed mood | ICD-10-CM |
| | F32.8 | Other depressive episodes | ICD-10-CM |
| | F32.9 | Major depressive disorder, single episode, unspecified | ICD-10-CM |
| | F32.A | Depression, unspecified | ICD-10-CM |
| | F32.89 | Other specified depressive episodes | ICD-10-CM |
| F41.0 | F41.0 | Panic disorder (episodic paroxysmal anxiety) | ICD-10-CM |
| | F41.8 | Other specified anxiety disorders | ICD-10-CM |
| | 300.02 | States, neurotic anxiety | ICD-9-CM |
| | F41.1 | Generalized anxiety disorder | ICD-10-CM |
| | F41.9 | Anxiety disorder, unspecified | ICD-10-CM |
| G47.0 | 780.52 | Wakefulness | ICD-9-CM |
| | G47.00 | Insomnia, unspecified | ICD-10-CM |
| G47.30 | G47.33 | Obstructive sleep apnea (adult) (pediatric) | ICD-10-CM |
| | G47.30 | Sleep apnea, unspecified | ICD-10-CM |
| | 327.23 | Upper airway resistance sleep apnea syndrome | ICD-9-CM |
| | 780.57 | Unspecified sleep apnea | ICD-9-CM |
| G89.18 | G89.18 | Other acute postprocedural pain | ICD-10-CM |
| | 338.18 | Postoperative pain NOS | ICD-9-CM |
| G89.29 | 338.4 | Pain syndrome, chronic | ICD-9-CM |
| | G89.29 | Other chronic pain | ICD-10-CM |
| | 338.29 | Pain, chronic | ICD-9-CM |
| | G89.4 | Chronic pain syndrome | ICD-10-CM |
| G93.40 | G93.41 | Metabolic encephalopathy | ICD-10-CM |
| | G93.40 | Encephalopathy, unspecified | ICD-10-CM |
| I10 | I11.0 | Hypertensive heart disease with heart failure (CMS/HHS-HCC) | ICD-10-CM |
| | I13.0 | Hypertensive heart and chronic kidney disease with heart failure and stage 1 through stage 4 chronic kidney disease, or unspecified chronic kidney disease (CMS/HHS-HCC) | ICD-10-CM |
| | I13.2 | Hypertensive heart and chronic kidney disease with heart failure and with stage 5 chronic kidney disease, or end stage renal disease (CMS/HHS-HCC) | ICD-10-CM |
| | I12.0 | Hypertensive chronic kidney disease with stage 5 chronic kidney disease or end stage renal disease (CMS/HHS-HCC) | ICD-10-CM |
| | 401.9 | White coat syndrome with hypertension | ICD-9-CM |
| | I10 | Essential (primary) hypertension | ICD-10-CM |
| I25.5 | 794.31 | ST-segment elevation | ICD-9-CM |
| | I25.5 | Ischemic cardiomyopathy | ICD-10-CM |
| I27.2 | I27.2 | Other secondary pulmonary hypertension | ICD-10-CM |
| | 416.8 | Secondary pulmonary hypertension | ICD-9-CM |
| | I27.20 | Pulmonary hypertension, unspecified (CMS/HHS-HCC) | ICD-10-CM |
| I47.2 | 427.1 | Wide-complex tachycardia | ICD-9-CM |
| | I47.2 | Ventricular tachycardia (CMS/HHS-HCC) | ICD-10-CM |
| I48.91 | I48.19 | Other persistent atrial fibrillation (CMS/HHS-HCC) | ICD-10-CM |
| | I48.1 | Persistent atrial fibrillation (CMS/HHS-HCC) | ICD-10-CM |

| | | | |
|---|---|---|---|
| | 427.31 | Rapid atrial fibrillation (CMS/HHS-HCC) | ICD-9-CM |
| | I48.91 | Unspecified atrial fibrillation (CMS/HHS-HCC) | ICD-10-CM |
| I50.20 | 428.23 | Heart failure, systolic, acute on chronic (CMS/HHS-HCC) | ICD-9-CM |
| | 428.22 | Heart failure, systolic, chronic (CMS/HHS-HCC) | ICD-9-CM |
| | I50.20 | Unspecified systolic (congestive) heart failure (CMS/HHS-HCC) | ICD-10-CM |
| | I50.22 | Chronic systolic (congestive) heart failure (CMS/HHS-HCC) | ICD-10-CM |
| | I50.23 | Acute on chronic systolic (congestive) heart failure (CMS/HHS-HCC) | ICD-10-CM |
| I50.32 | I50.32 | Chronic diastolic (congestive) heart failure (CMS/HHS-HCC) | ICD-10-CM |
| | I50.33 | Acute on chronic diastolic (congestive) heart failure (CMS/HHS-HCC) | ICD-10-CM |
| | 428.32 | Heart failure, diastolic, chronic (CMS/HHS-HCC) | ICD-9-CM |
| I50.9 | I50.9 | Heart failure, unspecified (CMS/HHS-HCC) | ICD-10-CM |
| | 428.9 | Weak heart (CMS/HHS-HCC) | ICD-9-CM |
| I95.9 | I95.89 | Other hypotension | ICD-10-CM |
| | I95.9 | Hypotension, unspecified | ICD-10-CM |
| | 458.9 | Unspecified hypotension | ICD-9-CM |
| | 458.8 | Other specified hypotension | ICD-9-CM |
| IMO0001 | IMO0001 | Reserved for non-ICD9 billable problem concepts | ICD-9-CM |
| | IMO0001 | Reserved for inherently not codable concepts without codable children | ICD-10-CM |
| | IMO0002 | Reserved for concepts with insufficient information to code with codable children | ICD-9-CM |
| | IMO0002 | Reserved for concepts with insufficient information to code with codable children | ICD-10-CM |
| J18.1 | J69.0 | Pneumonitis due to inhalation of food and vomit (CMS/HHS-HCC) | ICD-10-CM |
| | J18.9 | Pneumonia, unspecified organism | ICD-10-CM |
| | J18.1 | Lobar pneumonia, unspecified organism (CMS-HCC) | ICD-10-CM |
| | 486 | Water on the lung (HHS-HCC) | ICD-9-CM |
| J43.2 | J43.8 | Other emphysema (CMS/HHS-HCC) | ICD-10-CM |
| | J43.9 | Emphysema, unspecified (CMS/HHS-HCC) | ICD-10-CM |
| | J47.9 | Bronchiectasis, uncomplicated (CMS/HHS-HCC) | ICD-10-CM |
| | J43.2 | Centrilobular emphysema (CMS/HHS-HCC) | ICD-10-CM |
| J81.0 | J91.0 | Malignant pleural effusion (CMS-HCC) | ICD-10-CM |
| | J94.8 | Other specified pleural conditions | ICD-10-CM |
| | J81.0 | Acute pulmonary edema (CMS/HHS-HCC) | ICD-10-CM |
| | J90 | Pleural effusion, not elsewhere classified | ICD-10-CM |
| | 511.9 | Unspecified pleural effusion | ICD-9-CM |
| | J91.8 | Pleural effusion in other conditions classified elsewhere | ICD-10-CM |
| | J81.1 | Chronic pulmonary edema (HHS-HCC) | ICD-10-CM |
| J96.90 | J96.21 | Acute and chronic respiratory failure with hypoxia (CMS/HHS-HCC) | ICD-10-CM |
| | 518.81 | Type II respiratory failure (CMS/HHS-HCC) | ICD-9-CM |
| | 518.84 | Respiratory failure, acute and chronic (CMS/HHS-HCC) | ICD-9-CM |
| | J80 | Acute respiratory distress syndrome (CMS/HHS-HCC) | ICD-10-CM |
| | J95.821 | Acute postprocedural respiratory failure (CMS-HCC) | ICD-10-CM |
| | J96.91 | Respiratory failure, unspecified with hypoxia (CMS/HHS-HCC) | ICD-10-CM |
| | J96.00 | Acute respiratory failure, unspecified whether with hypoxia or hypercapnia (CMS/HHS-HCC) | ICD-10-CM |
| | J96.90 | Respiratory failure, unspecified, unspecified whether with hypoxia or hypercapnia (CMS/HHS-HCC) | ICD-10-CM |
| | J96.01 | Acute respiratory failure with hypoxia (CMS/HHS-HCC) | ICD-10-CM |
| | J96.02 | Acute respiratory failure with hypercapnia (CMS/HHS-HCC) | ICD-10-CM |
| | J96.22 | Acute and chronic respiratory failure with hypercapnia (CMS/HHS-HCC) | ICD-10-CM |
| K35.80 | 540.9 | Suppurative appendicitis | ICD-9-CM |
| | K35.80 | Unspecified acute appendicitis | ICD-10-CM |
| K42.9 | K42.9 | Umbilical hernia without obstruction or gangrene | ICD-10-CM |
| | 553.1 | Umbilical hernias | ICD-9-CM |
| K63.1 | 569.83 | Ulcer causing bleeding and hole in wall of stomach or small intestine (CMS/HHS-HCC) | ICD-9-CM |
| | K63.1 | Perforation of intestine (nontraumatic) (CMS/HHS-HCC) | ICD-10-CM |
| K63.2 | 569.81 | Small bowel fistula | ICD-9-CM |
| | K63.2 | Fistula of intestine | ICD-10-CM |
| K65.1 | 567.22 | Suprahepatic abscess (CMS/HHS-HCC) | ICD-9-CM |
| | K65.1 | Peritoneal abscess (CMS/HHS-HCC) | ICD-10-CM |
| K65.9 | K65.8 | Other peritonitis (CMS/HHS-HCC) | ICD-10-CM |
| | K65.9 | Peritonitis, unspecified (CMS/HHS-HCC) | ICD-10-CM |
| K72.0 | 570 | Yellow atrophy, acute | ICD-9-CM |
| | K72.00 | Acute and subacute hepatic failure without coma (HHS-HCC) | ICD-10-CM |
| K74.60 | K74.60 | Unspecified cirrhosis of liver (CMS/HHS-HCC) | ICD-10-CM |
| | 571.2 | Portal cirrhosis | ICD-9-CM |
| K76.0 | K76.0 | Fatty (change of) liver, not elsewhere classified | ICD-10-CM |
| | 571.8 | Steatosis, liver | ICD-9-CM |
| K76.89 | 573.9 | Unspecified disorder of liver | ICD-9-CM |

| | | | |
|---|---|---|---|
| | K76.89 | Other specified diseases of liver | ICD-10-CM |
| K83.0 | K83.0 | Cholangitis | ICD-10-CM |
| | 576.1 | Suppurative cholangitis (CMS-HCC) | ICD-9-CM |
| K85.9 | K85.8 | Other acute pancreatitis | ICD-10-CM |
| | K85.90 | Acute pancreatitis without necrosis or infection, unspecified (HHS-HCC) | ICD-10-CM |
| | K86.3 | Pseudocyst of pancreas (HHS-HCC) | ICD-10-CM |
| | K85.9 | Acute pancreatitis, unspecified | ICD-10-CM |
| K91.89 | 997.49 | Other digestive system complications | ICD-9-CM |
| | K91.89 | Other postprocedural complications and disorders of digestive system | ICD-10-CM |
| L02.211 | 682.2 | Trunk abscess | ICD-9-CM |
| | L02.211 | Cutaneous abscess of abdominal wall | ICD-10-CM |
| L08.9 | 682.9 | Superficial bacterial skin infection | ICD-9-CM |
| | L08.9 | Local infection of the skin and subcutaneous tissue, unspecified | ICD-10-CM |
| M79.89 | M79.89 | Other specified soft tissue disorders | ICD-10-CM |
| | 729.81 | Toe swelling | ICD-9-CM |
| N17.0 | 584.5 | Vasomotor nephropathy (CMS-HCC) | ICD-9-CM |
| | N17.0 | Acute kidney failure with tubular necrosis (CMS-HCC) | ICD-10-CM |
| N17.9 | 584.9 | Unspecified acute renal failure | ICD-9-CM |
| | N17.9 | Acute kidney failure, unspecified (CMS-HCC) | ICD-10-CM |
| N18.6 | N18.6 | End stage renal disease (CMS/HHS-HCC) | ICD-10-CM |
| | 585.6 | Type 1 diabetes mellitus with end-stage renal disease (ESRD) | ICD-9-CM |
| N18.9 | N18.9 | Chronic kidney disease, unspecified | ICD-10-CM |
| | 585.9 | Uremic, neuropathy | ICD-9-CM |
| N50.8 | N50.8 | Other specified disorders of male genital organs | ICD-10-CM |
| | N50.89 | Other specified disorders of the male genital organs | ICD-10-CM |
| N63 | N63.20 | Unspecified lump in the left breast, unspecified quadrant | ICD-10-CM |
| | N63.10 | Unspecified lump in the right breast, unspecified quadrant | ICD-10-CM |
| | N63 | Unspecified lump in breast | ICD-10-CM |
| R06.00 | 786.05 | Winded | ICD-9-CM |
| | R06.00 | Dyspnea, unspecified | ICD-10-CM |
| | R06.02 | Shortness of breath | ICD-10-CM |
| | R06.89 | Other abnormalities of breathing | ICD-10-CM |
| | 786.09 | Yawnings | ICD-9-CM |
| R09.02 | R09.02 | Hypoxemia | ICD-10-CM |
| | 799.02 | Severe insufficiency of oxygen in the blood | ICD-9-CM |
| R10.11 | 789.01 | Right upper quadrant pain | ICD-9-CM |
| | R10.11 | Right upper quadrant pain | ICD-10-CM |
| R13.12 | 787.22 | Transfer dysphagia | ICD-9-CM |
| | R13.12 | Dysphagia, oropharyngeal phase | ICD-10-CM |
| R14.3 | R14.3 | Flatulence | ICD-10-CM |
| | 787.3 | Wind symptom | ICD-9-CM |
| R17 | 782.4 | Yellowing of skin | ICD-9-CM |
| | R17 | Unspecified jaundice | ICD-10-CM |
| R18.8 | 789.59 | Urine in abdomen | ICD-9-CM |
| | R18.8 | Other ascites | ICD-10-CM |
| R40.0 | R41.82 | Altered mental status, unspecified | ICD-10-CM |
| | R40.0 | Somnolence | ICD-10-CM |
| | R40.4 | Transient alteration of awareness | ICD-10-CM |
| R41.0 | R41.0 | Disorientation, unspecified | ICD-10-CM |
| | 780.99 | Visuospatial agnosias | ICD-9-CM |
| R49.0 | R49.1 | Aphonia | ICD-10-CM |
| | 784.41 | Voice, loss | ICD-9-CM |
| R50.81 | 780.61 | Neutropenia with fever | ICD-9-CM |
| | R50.81 | Fever presenting with conditions classified elsewhere | ICD-10-CM |
| R57.9 | 785.59 | Vasomotor collapse | ICD-9-CM |
| | R57.9 | Shock, unspecified (CMS/HHS-HCC) | ICD-10-CM |
| R59.1 | R59.1 | Generalized enlarged lymph nodes | ICD-10-CM |
| | 785.6 | Viruses, Lymphadenopathy-Associated | ICD-9-CM |
| R60.9 | R60.9 | Edema, unspecified | ICD-10-CM |
| | 782.3 | Swollen | ICD-9-CM |
| R63.4 | 783.21 | Weightloss | ICD-9-CM |
| | R63.4 | Abnormal weight loss | ICD-10-CM |
| R65.10 | R65.10 | Systemic inflammatory response syndrome (sirs) of non-infectious origin without acute organ dysfunction (CMS/HHS-HCC) | ICD-10-CM |
| | 995.92 | Systemic inflammatory response syndrome due to infection, organ dysfun | ICD-9-CM |
| R65.21 | R65.21 | Severe sepsis with septic shock (CMS/HHS-HCC) | ICD-10-CM |

| | | | |
|---|---|---|---|
| | 785.52 | Shock, septic (CMS/HHS-HCC) | ICD-9-CM |
| R68.89 | 780.96 | Vague bodily discomfort | ICD-9-CM |
| | R68.89 | Other general symptoms and signs | ICD-10-CM |
| R74.8 | R74.8 | Abnormal levels of other serum enzymes | ICD-10-CM |
| | 790.5 | Serum lipase elevation | ICD-9-CM |
| R76.8 | R76.8 | Other specified abnormal immunological findings in serum | ICD-10-CM |
| | 795.79 | Serologic abnormality | ICD-9-CM |
| R78.81 | 790.7 | Unspecified bacteremia | ICD-9-CM |
| | R78.81 | Bacteremia | ICD-10-CM |
| R79.1 | 790.92 | Protime increased | ICD-9-CM |
| R79.1 | R79.1 | Abnormal coagulation profile | ICD-10-CM |
| R91.8 | 793.19 | Other nonspecific abnormal finding of lung field | ICD-9-CM |
| | R91.8 | Other nonspecific abnormal finding of lung field | ICD-10-CM |
| R94.31 | R94.31 | Abnormal electrocardiogram (ECG) (EKG) | ICD-10-CM |
| | 794.31 | ST-segment elevation | ICD-9-CM |
| T14.8 | T14.8 | Other injury of unspecified body region | ICD-10-CM |
| | 879.8 | Wounds, multiple | ICD-9-CM |
| | T14.8XXA | Other injury of unspecified body region, initial encounter | ICD-10-CM |
| T38.0X5A | E932.0 | Triamcinolone adverse reaction | ICD-9-CM |
| | T38.0X5A | Adverse effect of glucocorticoids and synthetic analogues, initial encounter | ICD-10-CM |
| Z00.6 | V70.7 | Examination of participant or control in clinical research | ICD-9-CM |
| | Z00.6 | Encounter for examination for normal comparison and control in clinical research program | ICD-10-CM |
| Z17.0 | Z17.0 | Estrogen receptor positive status (ER+) | ICD-10-CM |
| | V86.0 | Estrogen receptor positive status (ER+) | ICD-9-CM |
| Z17.1 | V86.1 | Estrogen receptor negative status (ER-) | ICD-9-CM |
| | Z17.1 | Estrogen receptor negative status (ER-) | ICD-10-CM |
| Z45.2 | Z45.2 | Encounter for adjustment and management of vascular access device | ICD-10-CM |
| | V58.81 | Visit for intravenous catheter placement | ICD-9-CM |
| Z51.5 | V66.7 | Terminal patient care | ICD-9-CM |
| | Z51.5 | Encounter for palliative care | ICD-10-CM |
| Z68.41 | V85.41 | Body mass index 40.0-44.9, adult (CMS/HHS-HCC) | ICD-9-CM |
| | Z68.41 | Body mass index (BMI) 40.0-44.9, adult (CMS/HHS-HCC) | ICD-10-CM |
| Z76.82 | V49.83 | Organ transplant candidate | ICD-9-CM |
| | Z76.82 | Awaiting organ transplant status | ICD-10-CM |
| Z79.01 | V58.61 | Use of blood thinner with INR goal of 2.5-3.5 | ICD-9-CM |
| | Z79.01 | Long term (current) use of anticoagulants | ICD-10-CM |
| Z79.2 | V58.62 | Prophylactic antibiotic | ICD-9-CM |
| | Z79.2 | Long term (current) use of antibiotics | ICD-10-CM |
| Z79.4 | V58.67 | Long-term insulin use in type 2 diabetes | ICD-9-CM |
| | Z79.4 | Long term (current) use of insulin (CMS/HHS-HCC) | ICD-10-CM |
| Z79.899 | Z79.899 | Other long term (current) drug therapy | ICD-10-CM |
| | V58.69 | Personal history of ongoing treatment with raloxifene | ICD-9-CM |
| Z86.19 | V12.09 | Varicella infection, resolved | ICD-9-CM |
| | Z86.19 | Personal history of other infectious and parasitic diseases | ICD-10-CM |
| Z86.711 | V12.55 | Personal history of pulmonary embolism | ICD-9-CM |
| | Z86.711 | Personal history of pulmonary embolism | ICD-10-CM |
| Z86.718 | V12.51 | Personal history of venous thrombosis and embolus | ICD-9-CM |
| | Z86.718 | Personal history of other venous thrombosis and embolism | ICD-10-CM |
| Z94.4 | V42.7 | Transplanted, liver | ICD-9-CM |
| | Z94.4 | Liver transplant status (CMS/HHS-HCC) | ICD-10-CM |
| Z95.2 | Z95.2 | Presence of prosthetic heart valve | ICD-10-CM |
| | V43.3 | Tricuspid valve replacement | ICD-9-CM |
| Z95.810 | Z95.810 | Presence of automatic (implantable) cardiac defibrillator | ICD-10-CM |
| | V45.02 | Status post internal cardiac defibrillator procedure | ICD-9-CM |
| Z95.811 | V43.21 | History of heart assist device (CMS/HHS-HCC) | ICD-9-CM |
| | Z95.811 | Presence of heart assist device (CMS/HHS-HCC) | ICD-10-CM |
| Z98.84 | V45.86 | Status post gastric bypass for obesity | ICD-9-CM |
| | Z98.84 | Bariatric surgery status | ICD-10-CM |
| Z98.89 | Z98.890 | Other specified postprocedural states | ICD-10-CM |
| | Z98.89 | Other specified postprocedural states | ICD-10-CM |
| Z99.11 | Z99.11 | Dependence on respirator (ventilator) status (CMS/HHS-HCC) | ICD-10-CM |
| | V46.11 | Ventilator dependent (CMS/HHS-HCC) | ICD-9-CM |
| Z99.2 | V56.0 | Visit for slow continuous ultrafiltration of kidneys | ICD-9-CM |
| | V45.11 | Status post peritoneal dialysis (CMS-HCC) | ICD-9-CM |
| | Z49.31 | slow continuous ultrafiltration of kidneys | ICD-9-CM |
| | Z99.2 | Dependence on renal dialysis (CMS-HCC) | ICD-10-CM |

| | | | |
|---|---|---|---|
| Z99.89 | V46.8 | Other machine dependence | ICD-9-CM |
| | Z99.89 | Dependence on other enabling machines and devices | ICD-10-CM |

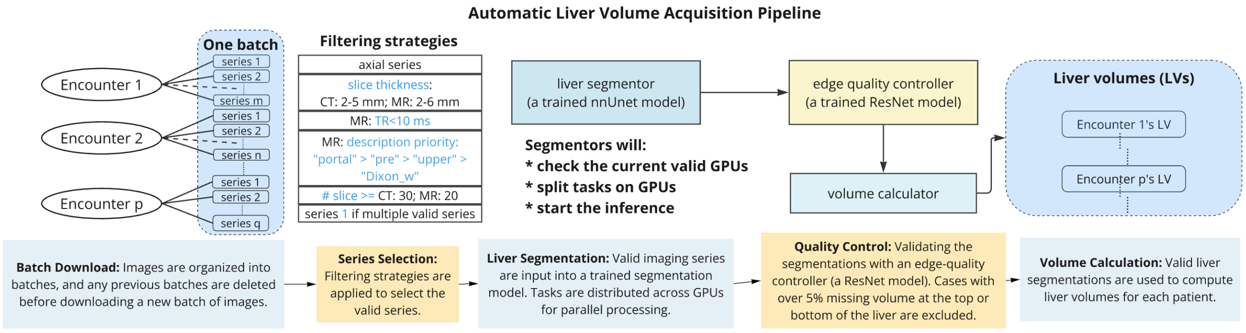


**Figure S1: Workflow diagram of automatic liver volume acquisition pipeline.** High-performance computing (HPC) clusters are used for segmentation and quality control, leading to the calculation of liver volumes for valid patient encounters.

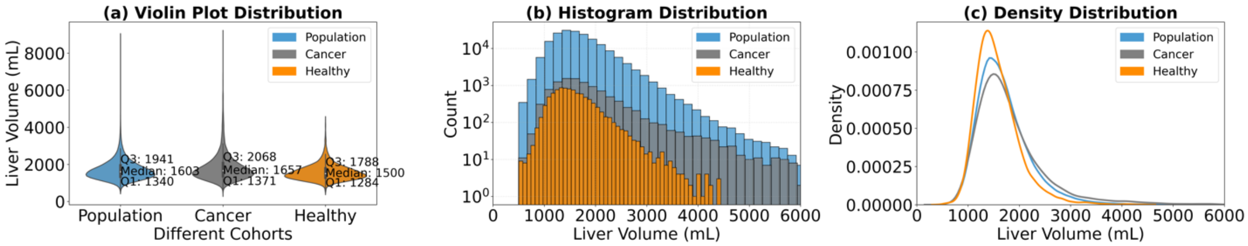


**Figure S2: Comparison of distribution statistics across three cohorts: Population, Cancer, and Healthy. (a)** Violin plots showing the distribution of values with medians and interquartile ranges for each group. **(b)** Log-scale histograms illustrating the frequency distribution of values across the groups. (c) Kernel density estimation (KDE) plots highlighting the probability density for each group.

### Section S1. Edge Quality Controller

For quality control, we implemented an edge-quality control mechanism using a ResNet-152 model. This classifier was trained to assess the completeness of the liver segmentation and exclude cases where more than 5% of the liver volume was missing from either the top or bottom of the organ. The training dataset was the same training set for the liver segmentor with complete liver in the scan, reported earlier in [20] and [21]. The complete dataset comprised 2,282 image-segmentation pairs, split into training, validation, and test sets at a 6:2:2 ratio.

The training data is generated by a three-channel contour overlap method from liver masks. The RGB channels of the input image represented the first slice containing the organ, the slice with the largest liver area, and the last slice containing the organ respectively. To simulate different levels of volume loss, we systematically removed a portion of the uppermost or lowermost slices to reflect realistic segmentation errors, as shown in Figure S3. By varying the number of removed slices, we created a range of cases with different degrees of truncation.

Using these preprocessed inputs, we trained a ResNet-152 model for binary classification, where class 0 indicates a segmentation below the quality threshold, and class 1 indicates a segmentation above the quality threshold. Binary Cross Entropy (BCE) loss was used for training, along with soft-thresholding and elastic regularization to enhance model robustness. The overall accuracy is 92% with false positive rate at 11.7% and false negative rate 7.2%. The trained model performance is shown in Table S2.

Table S2: Performance of the trained quality controller on the test set

| **Class** | **Precision** | **Recall** | **F1-score** |
|---|---|---|---|
| **0** (below threshold) | 0.77 | 0.88 | 0.82 |
| **1** (above threshold) | 0.97 | 0.93 | 0.95 |

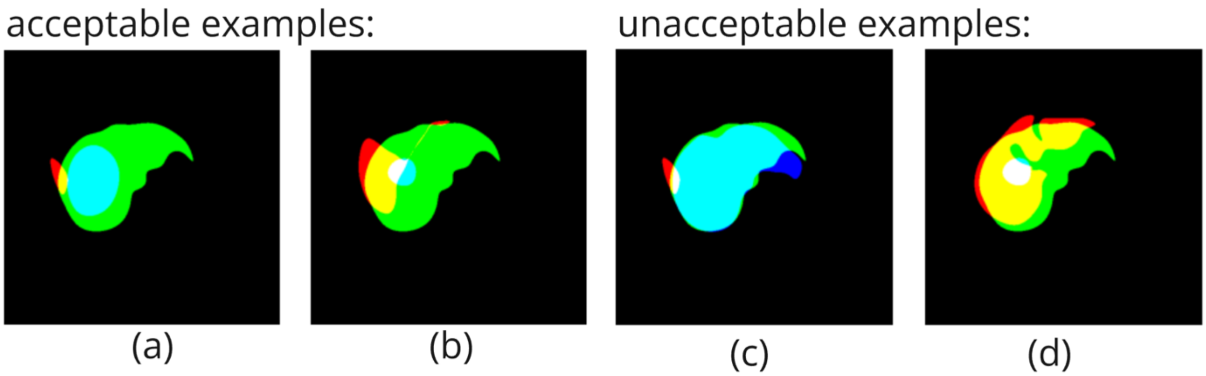


**Figure S3: Training data examples for liver segmentation quality control.**
